# Intrathoracic Adipose Tissue and Airway Structure, Lung Function, and Respiratory Symptoms: The Framingham Heart Study

**DOI:** 10.64898/2026.09.21.26363534

**Authors:** Jingzhou Zhang, Nicholas A. Bosch, Anica C. Law, Seung Hoan Choi, Kevin C. Wilson, Josée Dupuis, George R. Washko, Michael H. Cho, George T. O’Connor

**Affiliations:** The Pulmonary Center, Boston University Chobanian & Avedisian School of Medicine, Boston, Massachusetts, USA; Section of Pulmonary, Allergy, Sleep & Critical Care Medicine, Boston University Chobanian & Avedisian School of Medicine, Boston, Massachusetts, USA; Section of Preventive Medicine & Epidemiology, Boston University Chobanian & Avedisian School of Medicine, Boston, Massachusetts, USA; Department of Biostatistics, Boston University School of Public Health, Boston, Massachusetts, USA; Department of Epidemiology, Biostatistics and Occupational Health, McGill University, Montréal, Québec, Canada; Division of Pulmonary and Critical Care Medicine, Brigham and Women’s Hospital and Harvard Medical School, Boston, Massachusetts, USA; Channing Division of Network Medicine, Brigham and Women’s Hospital and Harvard Medical School, Boston, Massachusetts, USA; The NHLBI’s Framingham Heart Study, Framingham, Massachusetts, USA

**Keywords:** airway disease, asthma, COPD, intrathoracic fat, obesity, pericardial fat

## Abstract

**Background:** Obesity is associated with airway disease, but the contribution of specific thoracic adipose depots remains unclear. We examined whether intrathoracic adipose tissue (ITAT), defined as adipose tissue within the thoracic cavity excluding pericardial adipose tissue (PAT), was associated with airway phenotypes.

**Methods:** We included Framingham Heart Study participants with CT-derived thoracic adipose tissue measures. Cross-sectional associations of ITAT with CT-based airway wall and lumen measurements, spirometry, and respiratory symptoms were evaluated using multivariable regression. Primary analyses used sex-specific ITAT z-scores and adjusted for waist circumference to account for central obesity. Secondary analyses examined sex-specific ITAT quintiles and compared ITAT and PAT in individual-depot and mutually adjusted models.

**Results:** Among 2,825 adult participants (52.0% female), after adjustment for waist circumference, each 1-SD higher ITAT, relative to the sex-specific distribution, was associated with greater segmental (β [95% CI]: 0.010 mm [0.006, 0.014]) and subsegmental airway wall thickness (0.006 mm [0.002, 0.009]), narrower segmental airway lumen diameter (−0.049 [−0.088, −0.011]), lower FEV1 percent predicted (−1.92 percentage points [−2.72, −1.12]), and higher odds of wheezing (OR [95% CI]: 1.16 [1.02, 1.31]) and dyspnea (1.22 [1.05, 1.42]). Associations were similar after further adjustment for PAT and across ITAT quintiles. In contrast, PAT was not independently associated with airway structural measures after adjustment for ITAT.

**Conclusion:** Higher ITAT volume was associated with greater airway wall thickness and narrower airway lumen, as well as lower FEV_1_ and greater respiratory symptoms, independent of central obesity and pericardial adiposity. These findings support a depot-specific association of ITAT with airway disease.

**What is already known on this topic:** Obesity is associated with airway disease, but whether specific thoracic adipose depots have distinct associations with airway structure and function remains unclear.

**What this study adds:** In a community-based sample of adults, greater CT-derived intrathoracic adipose tissue volume, defined as adipose tissue within the thoracic cavity excluding pericardial adipose tissue, was associated with greater airway wall thickness, smaller airway lumen diameter, lower FEV_1_, and greater respiratory symptoms, independent of central obesity and pericardial adiposity.

**How this study might affect research, practice or policy:** These findings support a depot-specific association of intrathoracic adipose tissue with airway disease and warrant further investigation of its underlying biological mechanisms and potential relevance as a therapeutic target.

## Introduction

Excess adiposity, including overweight and obesity, is highly prevalent worldwide and contributes to cardiovascular disease and other medical conditions.^1–3^ Obesity has also long been associated with airway disease, such as asthma and phenotypes commonly observed in chronic obstructive pulmonary disease (COPD). For example, in patients with COPD, the “blue bloater” phenotype classically describes obese individuals with chronic bronchitis.^4^ In children, obesity is associated with airway dysanapsis, a mismatch between airway caliber and lung parenchymal size that predisposes to obstructive lung disease.^5,6^

Several mechanisms may underlie these associations. Central adiposity reduces lung compliance and promotes dynamic airway closure, leading to a restrictive pattern of lung function impairment with decreases in both FEV_1_ and FVC.^7,8^ Adipose tissue also functions as an endocrine organ that secretes adipokines and contributes to a chronic proinflammatory environment. For example, among individuals with asthma, higher body mass index (BMI) was associated with higher levels of inflammatory markers in blood and sputum.^9^ Using a polygenic score for BMI, it has recently been shown that genetic predisposition to higher BMI was associated with greater airway wall thickness, suggesting shared genetic underpinnings of obesity and bronchial wall thickening, a hallmark radiologic feature of airway disease.^10^

However, adipose deposits may not contribute equally to airway disease. Intrathoracic adipose tissue located within or adjacent to the airways may exert mechanical and paracrine influences on airways. Supporting this hypothesis, adipose tissue has been identified within large airways in postmortem lungs from persons with severe asthma and in porcine models, and it was associated with neutrophil and eosinophil infiltration and inflammatory markers in the airways.^11,12^ In addition, mediastinal fat has been associated with bronchial wall thickening in a COPD-enriched sample of male smokers.^13^ However, studies of patients with established COPD or asthma are susceptible to reverse causation bias, as disease severity and treatments such as corticosteroids may alter body composition.^14^ The contribution of airway-adjacent thoracic adipose tissue, independent of central adiposity and distinct from non-airway-adjacent thoracic fat depots, has not been well characterized in a large community-based population.

We hypothesized that intrathoracic adipose tissue (ITAT), comprising mediastinal and peribronchial fat, is associated with airway structure and pathophysiology beyond central obesity, represented by waist circumference, and non-airway-adjacent thoracic fat depot, represented by pericardial adipose tissue (PAT). To test this hypothesis, we examined associations between CT-derived ITAT volume and airway structural measures, as well as spirometry and respiratory symptoms, in a sample of community-dwelling adults.

## Methods

### Study population and design

We included participants from the Framingham Heart Study (FHS), an ongoing community-based cohort study that enrolled residents of Framingham, Massachusetts, United States.^15–17^ Briefly, the FHS was initiated in 1948 with the enrollment of 5,209 residents of Framingham. The study later expanded to include the Offspring cohort in 1971, comprising 5,124 children of the original participants and their spouses, and the Generation 3 cohort in 2002, consisting of 4,095 grandchildren. To enhance racial and ethnic diversity, the Omni 1 and Omni 2 cohorts were introduced in 1994 and 2003, enrolling 506 and 410 individuals, respectively, from underrepresented communities in Framingham and nearby areas. The FHS was approved by the Boston University Medical Campus Institutional Review Board, and all participants provided written informed consent.

Between 2008 and 2011, a total of 3,052 adult participants (men aged ≥35 years and women aged ≥40 years) from the Offspring, Generation 3, and Omni cohorts underwent whole-lung CT imaging using a GE Medical Systems VCT 64-slice PET/CT scanner. A study flow diagram of participant selection is provided in Figure 1.

**Figure 1.**
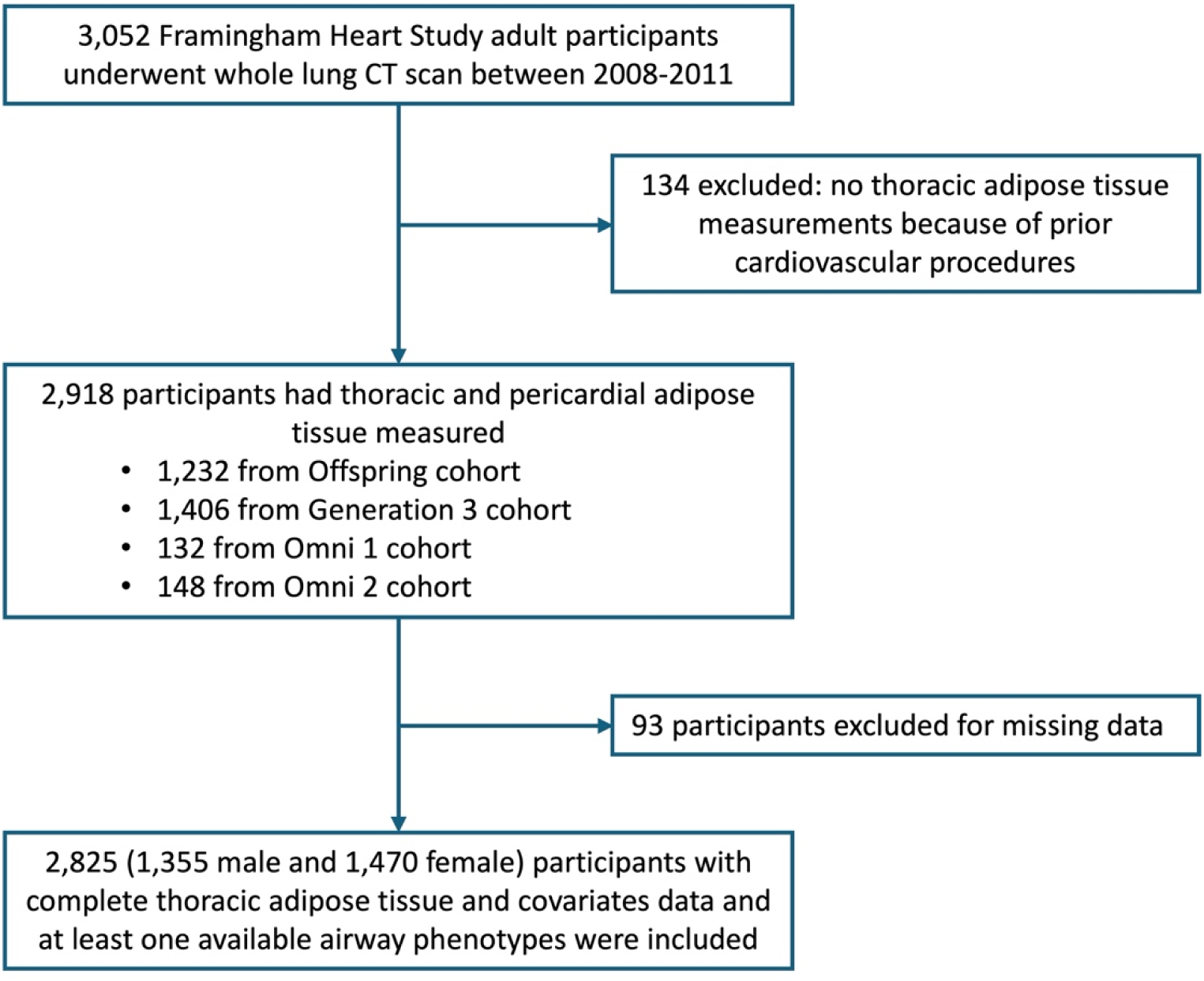
Flowchart of participant selection.

### Intrathoracic and pericardial adipose tissue measurements

Measurements of total thoracic and pericardial adipose tissue (PAT) volumes (cm^3^) have been previously described.^18^ Briefly, thoracic adipose tissue depots were quantified using a dedicated offline workstation (Aquarius 3D Workstation, TeraRecon Inc., San Mateo, CA) in 2,918 participants; 134 individuals were excluded due to prior cardiovascular procedures. A predefined display setting was used to identify adipose tissue, with a window width of −195 to −45 Hounsfield Units and a window center of −120 Hounsfield Units. Total thoracic adipose tissue and PAT volumes were quantified using a semi-automated segmentation approach. The reader manually traced the pericardium, after which segmentation across the imaging volume was automatically interpolated. Total thoracic adipose tissue was defined as all adipose tissue located within the thoracic cavity from the level of the right pulmonary artery to the diaphragm, and bounded by the chest wall and the descending aorta. PAT was defined as adipose tissue located within the pericardial sac. Intrathoracic adipose tissue (ITAT) volume was calculated as total thoracic adipose tissue volume minus PAT volume. This method demonstrated excellent reproducibility in prior studies, with intra-reader intraclass correlation coefficients of 0.99 for total thoracic adipose tissue and 0.97 for PAT, and inter-reader intraclass correlation coefficients of 0.98 for total thoracic adipose tissue and 0.95 for PAT.^18^

### Airway disease outcome measurements

Airway-related outcomes were grouped into three domains: 1) CT-based airway measures, including mean wall thickness and lumen diameter of segmental and subsegmental airways, to characterize airway wall thickening and luminal narrowing; 2) pre-bronchodilator spirometry measures, including percent predicted forced expiratory volume in 1 second (FEV_1_) and the ratio of FEV_1_ to forced vital capacity (FVC), to assess airflow limitation; and 3) self-reported respiratory symptoms, including wheeze, chronic cough, and dyspnea. Segmental and subsegmental airway wall thickness were prespecified as the primary outcomes, with the remaining measures considered secondary outcomes. This hierarchy was based on the hypothesis that airway thickening represents proximal manifestations of the association between ITAT and airway disease, with airway luminal narrowing, lung function impairment and respiratory symptoms representing downstream manifestations.

Airway measurements were extracted from CT images that were of sufficient quality for airway analysis (n = 2,501). Airway analysis was performed in the right upper and right lower lobes using software based on 3D Slicer (www.Slicer.org). Measurement sites were selected manually by visual inspection of the CT scans. At each selected site, segmental and subsegmental airway lumen and wall dimensions were measured. Airway assessment was performed across as many contiguous axial images within each bronchial segment of interest as possible, and the mean of these measurements was used for analysis.

Spirometry, respiratory questionnaires, and physical examination data were obtained from research visit examination cycles temporally closest to the CT scans used for ITAT and airway measurements, including Offspring exam 8, Generation 3 exam 2, Omni 1 exam 4, and Omni 2 exam 2. The median (Q1, Q3) interval between the CT scans and examination was 1.4 (0.1, 3.6) years. Wheezing was defined as a positive response to the question “wheezing or whistling in the chest in the past 12 months.” Chronic cough was defined as a positive response to the question “do you usually have a cough.” Dyspnea was defined as a modified Medical Research Council dyspnea scale of ≥1. Participants were included if they had at least one available airway-related outcome.

### Statistical analysis

Baseline characteristics were summarized using median (interquartile range [IQR]) for continuous variables and count (percentage) for categorical variables. We evaluated correlations of ITAT volume with BMI, waist circumference, and PAT volume using Spearman’s rank correlation coefficients. Given marked sex differences in ITAT distribution and body habitus, primary analyses used sex-specific z-scores of ITAT, PAT, and waist circumference to balance statistical power and interpretability. Associations of ITAT z-scores with airway outcomes were examined using mixed-effects models implemented with the glmmkin function in the GMMAT R package, accounting for known familial relationships. Logistic models were used for binary outcomes and Gaussian models for continuous outcomes. Primary models adjusted for waist circumference to account for central obesity, in addition to core covariates of age, current smoking, smoking pack-years, height, and study cohort. We conducted sensitivity analyses with additional adjustment for BMI as a marker of overall adiposity and for CT-derived lung volume as a marker of inspiratory effort at the time of CT acquisition to assess the robustness of the primary findings. BMI was not included in the primary models in addition to waist circumference because we considered central adiposity to be more relevant to airway disease, and because of the substantial correlation between these anthropometric measures. CT-derived lung volume was not included in the primary models because it was available only in a subset of participants and may partly reflect physiological consequences of obesity rather than act solely as a confounding factor. Missing data were handled using complete-case analysis for each outcome; therefore, sample sizes varied across outcomes according to availability of the specific airway phenotype.

To assess potential effect modification by sex, we tested ITAT-by-sex interaction terms and performed sex-stratified analyses. To facilitate clinical interpretations of ITAT’s associations with airway outcomes beyond a linear scale and assess for obvious nonlinear relationships, we conducted secondary analyses using sex-specific ITAT quintiles as the exposure, with the lowest quintile serving as the reference group. To compare the associations of ITAT and PAT with airway outcomes independent of central adiposity, we additionally fitted models examining PAT z-scores in relation to airway outcomes, adjusting for waist circumference and the same core covariates used in the primary analyses. To further evaluate depot-specific associations, we fitted mutually adjusted models that simultaneously included ITAT and PAT z-scores and examined changes in their effect estimates and statistical significance relative to the corresponding individual-depot models. All analyses were performed using R version 4.1. For the two prespecified primary outcomes of segmental and subsegmental airway wall thickness, a Bonferroni-adjusted two-sided significance threshold of *P* <0.025 was used. For secondary outcomes, a two-sided nominal *P* <0.05 was used.

## Results

### Study sample characteristics

Among 2,825 Framingham Heart Study participants predominantly of European ancestry, the median age was 56.0 years and 52.0% were female. Thoracic adipose tissue distribution differed substantially by sex, with median (IQR) ITAT volumes of 113.5 (88.8) cm^3^ in males and 54.9 (49.2) cm^3^ in females, and corresponding PAT volumes of 111.7 (61.9) cm^3^ and 89.2 (54.4) cm^3^, respectively. Smoking status distributions were similar between sexes, although females had lower cumulative smoking exposure than males. Compared with males, females on average had thinner wall and narrower lumen of airways, while FEV_1_ percent predicted and FEV_1_/FVC were similar between sexes. A higher proportion of females reported dyspnea, whereas the prevalence of wheezing and cough was similar between sexes. Additional study participants characteristics stratified by sex are shown in Table 1.

**Table 1.** Characteristics of study participants by sex.

|  | Male (n=1355) | Female (n=1470) |
| --- | --- | --- |
| Age (years) | 55.0 (16.0) | 58.0 (14.0) |
| Smoking status |  |  |
| Current smoker | 91 (6.7%) | 84 (5.7%) |
| Former smoker | 521 (38.5%) | 628 (42.7%) |
| Never smoker | 743 (54.8%) | 758 (51.6%) |
| Smoking pack-years * | 20.0 (23.1) | 14.5 (20.9) |
| Body mass index (kg/m<sup>2</sup>) | 28.2 (5.4) | 26.8 (7.8) |
| Waist circumference (cm) | 101.6 (14.0) | 95.3 (21.0) |
| Intrathoracic adipose tissue volume (cm<sup>3</sup>) | 113.5 (88.8) | 54.9 (49.2) |
| Pericardial adipose tissue volume (cm<sup>3</sup>) | 111.7 (61.9) | 89.2 (54.4) |
| Segmental airway measurements |  |  |
| Mean wall thickness (mm) | 1.10 (0.10) | 1.05 (0.10) |
| Mean lumen diameter (mm) | 5.0 (1.0) | 4.7 (0.9) |
| Subsegmental airway measurements |  |  |
| Mean wall thickness (mm) | 1.05 (0.10) | 1.00 (0.05) |
| Mean lumen diameter (mm) | 4.1 (1.0) | 3.7 (0.8) |
| FEV<sub>1</sub> percent predicted (%) | 104.8 (18.1) | 103.7 (20.9) |
| FEV<sub>1</sub>/FVC (%) | 76.0 (9.0) | 76.0 (8.0) |
| Respiratory symptoms |  |  |
| Wheezing | 246 (18.2%) | 260 (17.7%) |
| Chronic cough | 84 (6.2%) | 116 (7.9%) |
| Dyspnea | 85 (6.3%) | 201 (13.7%) |

\* Among ever-smokers
Continuous variables are presented as median (IQR) and categorical data are presented as count (%).
Of 1,355 male participants, 232 were missing segmental airway measurements, 253 were missing subsegmental airway measurements, and 260 were missing spirometry data for FEV<sub>1</sub> and FEV<sub>1</sub>/FVC. For respiratory symptom data, wheezing was missing in 3 participants, cough in 2 participants, and dyspnea in 9 participants.
Of 1,470 female participants, 230 were missing segmental airway measurements, 264 were missing subsegmental airway measurements, 171 were missing spirometry data for FEV<sub>1</sub> and FEV<sub>1</sub>/FVC, and 5 did not report dyspnea.
Wheezing was defined as present if a participant responded “yes” to the question “wheezing or whistling in the chest in the past 12 months.” Chronic cough was defined as present if a participant responded “yes” to the question “do you usually have cough.” Dyspnea was defined as present if a participant had a Modified Medical Research Council dyspnea scale of $\geq 1$ .
FEV<sub>1</sub>, forced expiratory volume in 1 second; FVC, forced vital capacity

### Correlation between ITAT and other adiposity measures

ITAT volume was positively correlated with BMI (Spearman correlation coefficient [95% CI], 0.58 [0.54, 0.62] in males and 0.60 [0.56, 0.63] in females), waist circumference (0.67 [0.64, 0.70]; 0.66 [0.63, 0.69]), and PAT volume (0.72 [0.69, 0.75]; 0.72 [0.69, 0.74]). Correlations were strongest for PAT volume and were broadly similar between sexes.

### Associations between ITAT and airway outcomes

Results from multivariable regression models examining the associations of ITAT with airway-related outcomes are shown in Table 2. After adjustment for waist circumference, a 1-standard-deviation (SD) higher ITAT volume, relative to the sex-specific distribution, was associated with greater mean wall thickness in both segmental (β [95% CI], 0.010 mm [0.006, 0.014]) and subsegmental airways (0.006 mm [0.002, 0.009]) and with smaller segmental airway lumen diameter (−0.049 mm [−0.088, −0.011]), but was not associated with subsegmental airway lumen diameter. Higher ITAT volume was also associated with lower FEV_1_ percent predicted (−1.92% [- 2.72, −1.12]) and higher odds of wheeze (OR [95% CI], 1.16 [1.02, 1.31]) and dyspnea (1.22 [1.06, 1.42]), but not chronic cough.

**Table 2.**
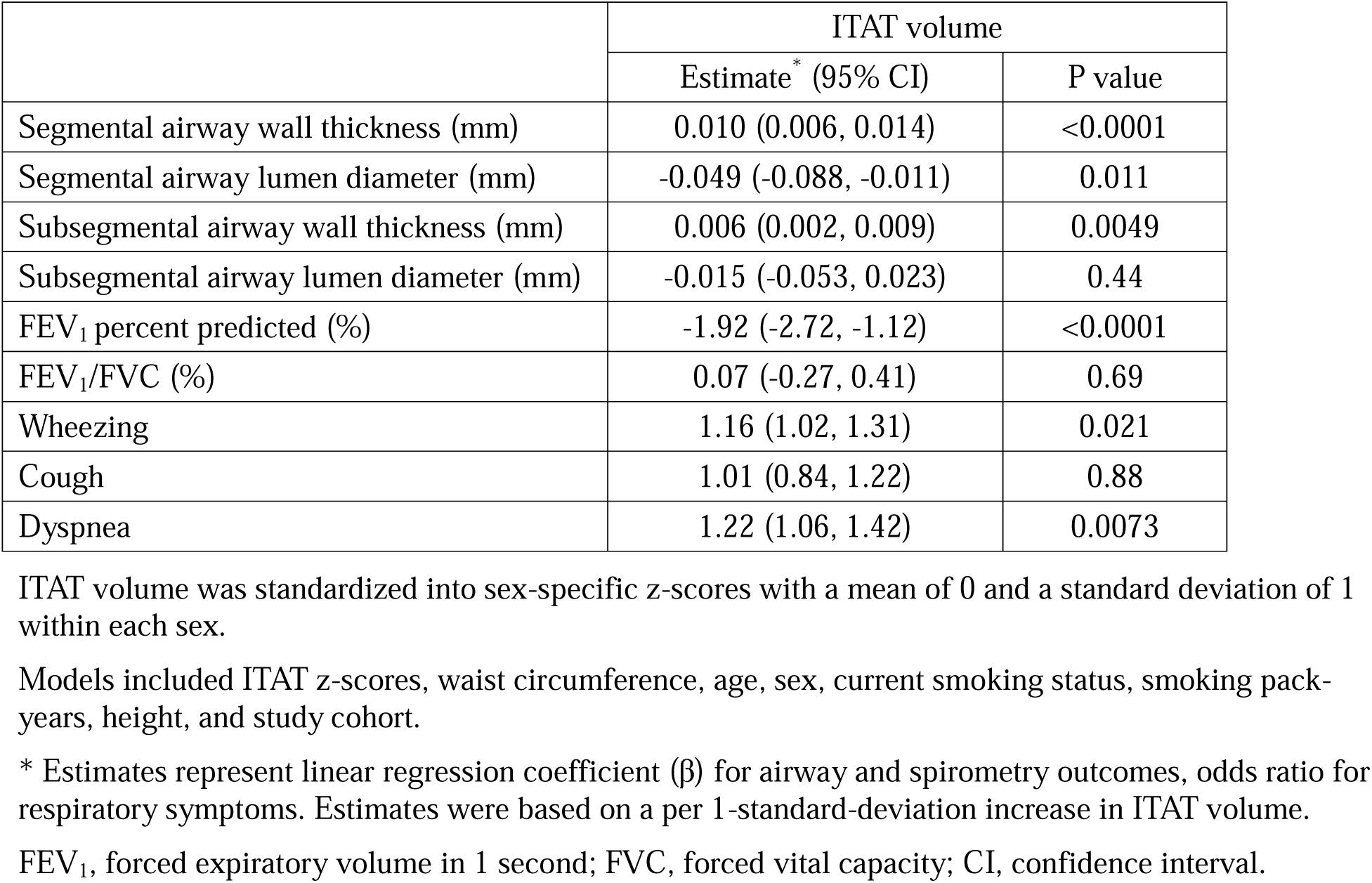
Multivariable regression analyses of the associations between intrathoracic adipose tissue (ITAT) and airway-related outcomes.

|  | ITAT volume |  |
| --- | --- | --- |
|  | Estimate* (95% CI) | P value |
| Segmental airway wall thickness (mm) | 0.010 (0.006, 0.014) | <0.0001 |
| Segmental airway lumen diameter (mm) | -0.049 (-0.088, -0.011) | 0.011 |
| Subsegmental airway wall thickness (mm) | 0.006 (0.002, 0.009) | 0.0049 |
| Subsegmental airway lumen diameter (mm) | -0.015 (-0.053, 0.023) | 0.44 |
| FEV<sub>1</sub> percent predicted (%) | -1.92 (-2.72, -1.12) | <0.0001 |
| FEV<sub>1</sub>/FVC (%) | 0.07 (-0.27, 0.41) | 0.69 |
| Wheezing | 1.16 (1.02, 1.31) | 0.021 |
| Cough | 1.01 (0.84, 1.22) | 0.88 |
| Dyspnea | 1.22 (1.06, 1.42) | 0.0073 |

ITAT volume was standardized into sex-specific z-scores with a mean of 0 and a standard deviation of 1 within each sex.
Models included ITAT z-scores, waist circumference, age, sex, current smoking status, smoking pack-years, height, and study cohort.
\* Estimates represent linear regression coefficient ( $\beta$ ) for airway and spirometry outcomes, odds ratio for respiratory symptoms. Estimates were based on a per 1-standard-deviation increase in ITAT volume.
FEV<sub>1</sub>, forced expiratory volume in 1 second; FVC, forced vital capacity; CI, confidence interval.

To contextualize the magnitude of associations between ITAT and airway measurements, each 10-pack-year increase in cigarette smoking was associated with thicker segmental (0.002 mm [0.0003, 0.004]) and subsegmental (0.002 mm [0.0002, 0.004]) airway walls and narrower segmental (−0.020 mm [−0.040, −0.0001]) and subsegmental (−0.028 mm [−0.048, −0.008]) lumen diameters.

In analyses additionally adjusted for BMI, the associations between ITAT and airway outcomes were essentially unchanged (Supplementary Table S1). Among participants with available CT-derived lung volume data, further adjustment for lung volume modestly attenuated several associations, but the overall pattern of findings remained consistent (Supplementary Table S2).

### Effect modification by sex and sex-stratified analysis

In models including an ITAT-by-sex interaction term, with males as the reference group, we did not observe a consistent pattern of effect modification by sex across airway outcomes. Nominal evidence of interaction was observed for subsegmental airway wall thickness (interaction β =-0.007 mm, *P*=0.018), suggesting that the increase in subsegmental airway wall thickness associated with a 1-SD higher ITAT volume was 0.007 mm smaller in females than in males (Supplementary Table S3). Because ITAT was standardized within sex, interaction terms should be interpreted as sex differences in associations per 1-SD higher ITAT within the sex-specific distribution, rather than per absolute ITAT volume.

In sex-stratified analyses (Supplementary Table S4), numerically stronger associations of ITAT with subsegmental airway wall thickness (β [95% CI], 0.009 mm [0.003, 0.015] vs. 0.002 mm [−0.003, 0.007]) and FEV_1_ percent predicted (−2.80% [−3.99, −1.60] vs. −1.20% [−2.29, −0.12]) were observed in males than in females. However, the overall direction and pattern of associations between ITAT and airway outcomes were generally consistent across sexes.

### ITAT quintiles and airway outcomes

As shown in Figure 2, increasing ITAT quintiles were associated with progressively thicker airway walls, lower FEV1 percent predicted, and higher odds of wheezing and dyspnea, consistent with a dose-response relationship. Compared with participants in the lowest quintile (Q1), those in the highest quintile (Q5) had 0.026 mm (95% CI: 0.014, 0.038) greater segmental airway wall thickness, 0.014 mm (0.002, 0.025) greater subsegmental airway wall thickness, 4.80% (2.38, 7.21) lower FEV_1_ percent predicted, and higher odds of wheezing (OR [95% CI]: 2.06 [1.34, 3.15]) and dyspnea (1.83 [1.03, 3.24]) (Supplementary Table S5). While certain higher ITAT quintiles were associated with lower segmental airway lumen diameter and FEV_1_/FVC compared with Q1, no clear dose-response relationship was observed, which may reflect a nonlinear association, threshold effects, or limited precision of the quintile estimates.

**Figure 2.**
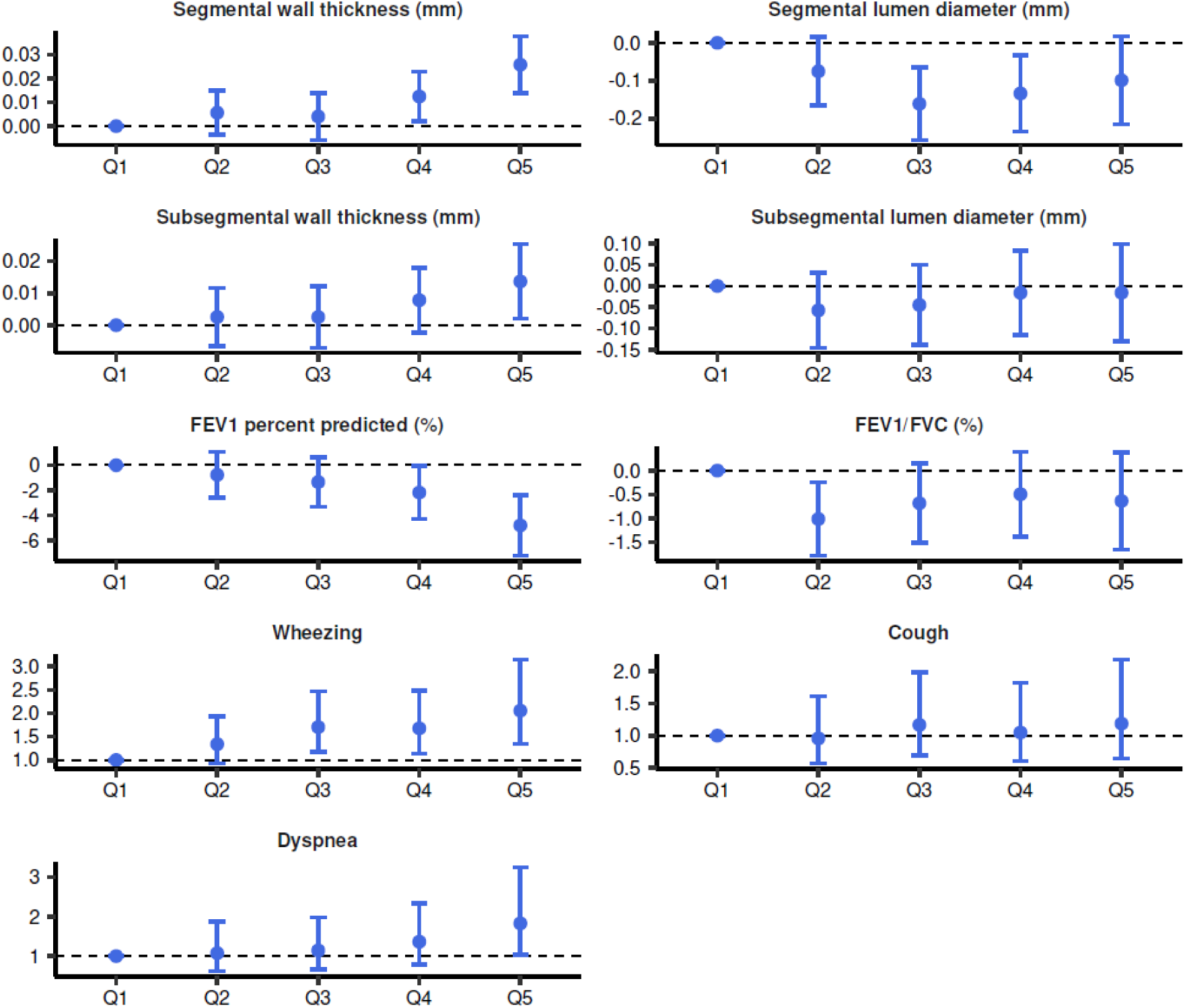
Association between intrathoracic adipose tissue (ITAT) volume quintiles and airway disease-related phenotypes. Models included sex-specific ITAT quintiles, waist circumference, age, sex, current smoking status, smoking pack-years, height, and study cohort.

### Comparison of ITAT and PAT

Results from multivariable regression models examining associations between PAT and airway outcomes are shown in Table 3. After adjustment for waist circumference, higher PAT was associated with greater segmental airway wall thickness but not with other airway measures, as well as higher FEV_1_/FVC and greater odds of cough.

**Table 3.** Multivariable regression analyses of the associations between pericardial adipose tissue (PAT) and airway-related outcomes.

|  | PAT volume |  |
| --- | --- | --- |
|  | Estimate* (95% CI) | P value |
| Segmental airway wall thickness (mm) | 0.005 (0.001, 0.008) | 0.012 |
| Segmental airway lumen diameter (mm) | -0.032 (-0.069, 0.004) | 0.083 |
| Subsegmental airway wall thickness (mm) | 0.002 (-0.002, 0.005) | 0.32 |
| Subsegmental airway lumen diameter (mm) | -0.005 (-0.041, 0.031) | 0.79 |
| FEV<sub>1</sub> percent predicted (%) | -0.27 (-1.03, 0.50) | 0.50 |
| FEV<sub>1</sub>/FVC (%) | 0.42 (0.10, 0.73) | 0.011 |
| Wheezing | 1.05 (0.93, 1.19) | 0.41 |
| Cough | 1.29 (1.09, 1.53) | 0.0026 |
| Dyspnea | 1.08 (0.93, 1.25) | 0.31 |

PAT volume was standardized into sex-specific z-scores with a mean of 0 and a standard deviation of 1 within each sex.
Models included PAT z-scores, waist circumference, age, sex, current smoking status, smoking pack-years, height, and study cohort.
\* Estimates represent linear regression coefficient ( $\beta$ ) for airway and spirometry outcomes, odds ratio for respiratory symptoms. Estimates were based on a per 1-standard-deviation increase in PAT volume.
FEV<sub>1</sub>, forced expiratory volume in 1 second; FVC, forced vital capacity; CI, confidence interval.

In models jointly including ITAT and PAT (Table 4), ITAT effect estimates were generally unchanged after adjustment for PAT, and associations with airway wall thickness, FEV_1_ percent predicted, wheeze, and dyspnea remained significant. In contrast, PAT associations with airway structural measures were attenuated after adjustment for ITAT, whereas associations with FEV_1_/FVC and cough persisted. PAT also showed a nominal positive association with FEV_1_ percent predicted only in the mutually adjusted model. Overall, these findings support more ITAT-specific associations with airway structure and distinct depot-specific associations with lung function and respiratory symptoms.

**Table 4.** Associations of intrathoracic and pericardial adipose tissue volumes with airway-related outcomes in mutually adjusted models.

|  | ITAT volume |  | PAT volume |  |
| --- | --- | --- | --- | --- |
|  | Estimate* (95% CI) | P value | Estimate* (95% CI) | P value |
| Segmental airway wall thickness (mm) | 0.010 (0.005, 0.014) | <0.0001 | 0.001 (-0.004, 0.005) | 0.81 |
| Segmental airway lumen diameter (mm) | -0.044 (-0.089, 0.001) | 0.054 | -0.010 (-0.052, 0.032) | 0.64 |
| Subsegmental airway wall thickness (mm) | 0.006 (0.001, 0.0010) | 0.014 | -0.000 (-0.004, 0.004) | 0.93 |
| Subsegmental airway lumen diameter (mm) | -0.017 (-0.061, 0.028) | 0.46 | 0.003 (-0.038, 0.044) | 0.89 |
| FEV<sub>1</sub> percent predicted (%) | -2.50 (-3.44, -1.57) | <0.0001 | 1.06 (0.17, 1.95) | 0.020 |
| FEV<sub>1</sub>/FVC (%) | -0.27 (-0.66, 0.13) | 0.18 | 0.61 (0.24, 0.98) | 0.0013 |
| Wheezing | 1.17 (1.01, 1.36) | 0.033 | 0.98 (0.85, 1.13) | 0.77 |
| Cough | 0.85 (0.68, 1.06) | 0.15 | 1.37 (1.12, 1.66) | 0.0017 |
| Dyspnea | 1.25 (1.05, 1.49) | 0.013 | 0.96 (0.81, 1.14) | 0.67 |

ITAT and PAT volumes were standardized into sex-specific z-scores with a mean of 0 and a standard deviation of 1 within each sex.
Models included ITAT z-scores, PAT z-scores, age, sex, current smoking status, smoking pack-years, height, and study cohort.
\* Estimates represent linear regression coefficient ( $\beta$ ) for airway and spirometry outcomes, odds ratio for respiratory symptoms. Estimates were based on per 1-standard-deviation increase in ITAT volume or waist circumference.
ITAT, intrathoracic adipose tissue; PAT, pericardial adipose tissue; FEV<sub>1</sub>, forced expiratory volume in 1 second; FVC, forced vital capacity; CI, confidence interval.

## Discussion

In this community-based sample of adults, CT-derived intrathoracic adipose tissue (ITAT), defined as adipose tissue within the thoracic cavity excluding pericardial adipose tissue (PAT), was associated with multiple airway-related phenotypes, including CT-based airway structural measures, spirometry measures, and respiratory symptoms, after adjustment for waist circumference. These associations were also observed after further adjustment for PAT, supporting an ITAT-specific association with airway disease manifestations beyond central and pericardial adiposity.

A central question in obesity-related airway disease is whether excess adiposity primarily reflects generalized body habitus and restrictive respiratory mechanics or whether specific fat depots have more direct relevance to airway pathology. Airway wall thickening is a key CT manifestation of airway inflammation and remodeling and has been associated with respiratory symptoms and adverse clinical outcomes in individuals with and without obstructive airway disease.^19–21^ In our analysis, higher ITAT volume was associated with greater segmental and subsegmental airway wall thickness independent of central adiposity and PAT, whereas PAT showed no independent association with airway structural measures after accounting for ITAT. Although ITAT represents a heterogeneous mixture of thoracic fat depots, including mediastinal, peribronchial, and perivascular adipose tissues, and density-based CT quantification may not precisely capture the relatively small amount of peribronchial fat, the contrasting findings for ITAT and the non-airway-adjacent PAT depot support a more depot-specific association of ITAT with airway structure. Given the anatomical proximity of components of ITAT to the airways, these findings raise the possibility that local thoracic adipose tissue may contribute to airway inflammation and remodeling.

The spirometry findings further distinguished ITAT from central adiposity and PAT. In waist circumference-adjusted models, higher ITAT was associated with lower FEV_1_, whereas its association with FEV_1_/FVC was inconsistent, with no significant association in the continuous analysis and lower FEV_1_/FVC observed only in selected ITAT quintiles. In contrast, greater waist circumference and PAT were associated with higher FEV_1_/FVC. Together, these findings suggest that ITAT is primarily associated with reduced FEV_1_ without a consistent reduction in FEV_1_/FVC, a pattern potentially compatible with preserved ratio impaired spirometry rather than classic obstructive physiology. This distinction may be clinically relevant given that preserved ratio impaired spirometry (PRISm) is associated with increased risk of subsequent airflow obstruction and adverse clinical outcomes. Given the unmet clinical need for effective therapies for PRISm, further studies are warranted to determine whether ITAT may represent a potentially modifiable or treatable trait.

In the Framingham Heart Study, PAT volume has previously been associated with cardiac structure and cardiovascular diseases, including myocardial infarction and atrial fibrillation.^22–24^ Despite similar adipose tissue volumes, ITAT and PAT showed distinct associations with airway-related outcomes. In contrast to ITAT, PAT was not independently associated with airway structural measures. Instead, higher PAT was associated with higher FEV_1_/FVC, a pattern potentially consistent with restrictive physiology and consistent with findings from the Jackson Heart Study.^25^ The positive association between PAT and FEV_1_ emerged only after mutual adjustment for ITAT and should therefore be interpreted cautiously; this pattern may reflect a suppression effect related to the positive correlation between the two thoracic fat depots and their divergent associations with lung function. For respiratory symptoms, ITAT was associated with wheeze and dyspnea, whereas PAT was associated with cough. Wheeze and dyspnea may be related to the airway wall thickening and reduced FEV_1_ associated with ITAT, whereas the mechanism underlying the association between PAT and cough is less clear. Given the absence of independent associations between PAT and airway structural measures, this relationship may involve non-airway pathways, including cardiometabolic comorbidities associated with PAT. Together, these findings support the concept that the anatomical distribution of thoracic adipose tissue may influence its associations with cardiopulmonary phenotypes.

These findings may also have clinical implications. Randomized controlled trials have shown that weight management can improve lung function and clinical outcomes in COPD and asthma,^26–28^ supporting obesity as a treatable trait in airway disease. In our study, a 1-SD increase in ITAT volume was associated with segmental airway wall thickening comparable in magnitude to that associated with approximately 40 pack-years of smoking, a 1.9-percentage-point lower FEV_1_ percent predicted, and approximately 20% higher odds of wheezing and dyspnea, independent of central obesity. These findings suggest that ITAT may represent a clinically relevant adipose depot. Chest CT is increasingly recognized as a valuable modality for the evaluation of airway disease, and CT-detected airway wall thickening and mucus plugging have been associated with disease severity and outcomes in COPD and asthma.^29–31^ Because intrathoracic adipose tissue can be quantified simultaneously on chest CT, it may provide additional information for airway disease evaluation. This possibility may be particularly relevant in the setting of emerging pharmacologic weight-management strategies, including glucagon-like peptide-1 (GLP-1) and dual GLP-1/gastric inhibitory polypeptide (GIP) agonists, which are increasingly being studied in patients with obesity and asthma.^32,33^ Evaluating changes in ITAT, in addition to other adipose depots, in such studies may help determine whether airway-adjacent thoracic adiposity contributes to treatment response and should be considered in selecting patients for therapy.

Strengths of this study include the large, community-based sample with balanced sex distribution; adjustment for central adiposity and pericardial fat to better isolate the association of ITAT with airway-related outcomes; and evaluation across structural, functional, and symptomatic domains. Consistent dose-response relationships across multiple outcomes also support the robustness and internal consistency of the findings.

This study has several limitations. First, although we inferred independent associations of ITAT and PAT with airway-related outcomes by jointly interpreting findings from the individual and mutually adjusted models, these results should be interpreted with caution given the substantial correlation between the two fat depots. Second, ITAT was quantified using CT density thresholds, and we were unable to distinguish specific intrathoracic fat compartments or to assess adipose tissue quality beyond fat volume. Third, airway wall measurements were derived from the right lung rather than averaged across all lobes. However, substantial asymmetric lung disease is unlikely in this community-based cohort, and any resulting measurement error would likely be nondifferential with respect to ITAT. Fourth, external replication is currently not available, and the generalizability of our findings to populations of non-European ancestry and outside the United States remains to be established. Finally, although we examined a broad range of airway-related outcomes, measures of small airway disease and longitudinal changes were not evaluated.

In conclusion, in a community-based sample, higher intrathoracic adipose tissue volume was associated with airway wall thickening and luminal narrowing, independent of central obesity and pericardial adiposity, and was also associated with reduced FEV_1_ and wheezing and dyspnea. In contrast, pericardial adipose tissue showed no independent association with airway structural measures. These findings support a depot-specific association of intrathoracic adipose tissue with airway disease and warrant further investigation into its underlying biological mechanisms and potential relevance as a therapeutic target.

## Data Availability

The Framingham Heart Study data used in this study are available through the National Center for Biotechnology Information Database of Genotypes and Phenotypes (dbGaP), subject to applicable data access requirements and approvals.

## Authors’ contributions

JZ conceptualized and designed the study and performed data analysis. All authors contributed to data interpretation. JZ drafted the manuscript, and all authors critically revised the manuscript and approved its submission. MHC and GTO jointly supervised this work.

## Disclosures

M.H.C. has received grant support from Bayer and Genentech, and consulting fees from Apogee Therapeutics and BMS. KCW is the Executive Vice-President of Guidelines and Documents at the American Thoracic Society. GRW has received consulting fees from 35Pharma, Agenx, AstraZeneca, Genentech, GlaxoSmithKline, Insmed, Merck, Montai Therapeutics, PulmonX, Rage Biotech, Regeneron, Sanofi, travel stipend from AstraZeneca, and is a co-founder and equity share holder in Quantitative Imaging Solutions. All other authors declared no conflict of interest.

A large language model (ChatGPT) was used to assist with language editing. The authors reviewed and edited the content as needed and take full responsibility for the content of the publication.

---

**Table S1.** Multivariable regression analyses of the associations between intrathoracic adipose tissue (ITAT) and airway-related outcomes, with additional adjustment for body mass index (BMI).

|  | ITAT volume |  |
| --- | --- | --- |
|  | Estimate* (95% CI) | P value |
| Segmental airway wall thickness (mm) | 0.010 (0.006, 0.014) | <0.0001 |
| Segmental airway lumen diameter (mm) | -0.051 (-0.090, -0.013) | 0.0088 |
| Subsegmental airway wall thickness (mm) | 0.005 (0.001, 0.009) | 0.013 |
| Subsegmental airway lumen diameter (mm) | -0.015 (-0.053, 0.023) | 0.44 |
| FEV<sub>1</sub> percent predicted (%) | -2.10 (-2.90, -1.29) | <0.0001 |
| FEV<sub>1</sub>/FVC (%) | -0.03 (-0.36, 0.31) | 0.88 |
| Wheezing | 1.15 (1.02, 1.31) | 0.026 |
| Cough | 1.04 (0.86, 1.25) | 0.68 |
| Dyspnea | 1.22 (1.05, 1.41) | 0.0095 |

ITAT volume was standardized into sex-specific z-scores with a mean of 0 and a standard deviation of 1 within each sex.
Models included ITAT z-scores, waist circumference, BMI, age, sex, current smoking status, smoking pack-years, height, and study cohort.
\* Estimates represent linear regression coefficient ( $\beta$ ) for airway and spirometry outcomes, odds ratio for respiratory symptoms. Estimates were based on a per 1-standard-deviation increase in ITAT volume.
FEV<sub>1</sub>, forced expiratory volume in 1 second; FVC, forced vital capacity; CI, confidence interval.

**Table S2.** Multivariable regression analyses of the associations between intrathoracic adipose tissue (ITAT) and airway-related outcomes, with additional adjustment for CT-derived lung volume.

|  | ITAT volume |  |
| --- | --- | --- |
|  | Estimate* (95% CI) | P value |
| Segmental airway wall thickness (mm) | 0.008 (0.004, 0.012) | <0.0001 |
| Segmental airway lumen diameter (mm) | -0.033 (-0.072, 0.005) | 0.092 |
| Subsegmental airway wall thickness (mm) | 0.004 (-0.000, 0.008) | 0.064 |
| Subsegmental airway lumen diameter (mm) | 0.011 (-0.028, 0.050) | 0.60 |
| FEV<sub>1</sub> percent predicted (%) | -1.83 (-2.69, -0.98) | <0.0001 |
| FEV<sub>1</sub>/FVC (%) | -0.26 (-0.63, 0.10) | 0.16 |
| Wheezing | 1.19 (1.04, 1.37) | 0.012 |
| Cough | 1.10 (0.90, 1.35) | 0.33 |
| Dyspnea | 1.15 (0.98, 1.36) | 0.086 |

ITAT volume was standardized into sex-specific z-scores with a mean of 0 and a standard deviation of 1 within each sex.
Models included ITAT z-scores, waist circumference, age, sex, current smoking status, smoking pack-years, height, CT-derived lung volume and study cohort.
\* Estimates represent linear regression coefficient ( $\beta$ ) for airway and spirometry outcomes, odds ratio for respiratory symptoms. Estimates were based on a per 1-standard-deviation increase in ITAT volume.
FEV<sub>1</sub>, forced expiratory volume in 1 second; FVC, forced vital capacity; CI, confidence interval.

**Table S3.** Interaction between intrathoracic adipose tissue (ITAT) volume and sex in models of airway outcomes adjusting for waist circumference.

|  | ITAT volume |  | Sex (female) |  | ITAT-by-sex |  |
| --- | --- | --- | --- | --- | --- | --- |
|  | Estimate | P value | Estimate | P value | Estimate | P value |
| Segmental airway wall thickness (mm) | $\beta = 0.013$ | <0.0001 | $\beta = -0.050$ | <0.0001 | $\beta = -0.005$ | 0.083 |
| Segmental airway lumen diameter (mm) | $\beta = -0.046$ | 0.063 | $\beta = -0.175$ | <0.0001 | $\beta = -0.007$ | 0.81 |
| Subsegmental airway wall thickness (mm) | $\beta = 0.009$ | 0.0002 | $\beta = -0.026$ | <0.0001 | $\beta = -0.007$ | 0.018 |
| Subsegmental airway lumen diameter (mm) | $\beta = -0.005$ | 0.83 | $\beta = -0.146$ | 0.0006 | $\beta = -0.020$ | 0.49 |
| FEV<sub>1</sub> percent predicted (%) | $\beta = -2.45$ | <0.0001 | $\beta = -4.07$ | <0.0001 | $\beta = 0.98$ | 0.10 |
| FEV<sub>1</sub>/FVC (%) | $\beta = 0.14$ | 0.53 | $\beta = -1.38$ | 0.0003 | $\beta = -0.13$ | 0.61 |
| Wheezing | OR = 1.16 | 0.066 | OR = 1.02 | 0.88 | OR = 1.00 | 1.0 |
| Cough | OR = 0.98 | 0.88 | OR = 1.24 | 0.34 | OR = 1.06 | 0.68 |
| Dyspnea | OR = 1.29 | 0.018 | OR = 1.56 | 0.042 | OR = 0.91 | 0.48 |

ITAT volume was standardized into z-scores prior to analysis. Models included ITAT, sex (male as reference), an ITAT × sex interaction term, waist circumference, age, current smoking status, smoking pack-years, height, and study cohort.
Estimate was based on per 1-standard-deviation increase in ITAT volume.
FEV<sub>1</sub>, forced expiratory volume in 1 second; FVC, forced vital capacity.; OR, odds ratio.

**Table S4.** Sex-stratified multivariable regression analyses of the associations between intrathoracic adipose tissue (ITAT) and airway-related outcomes.

| Outcome | ITAT volume |  |  |  |
| --- | --- | --- | --- | --- |
|  | Male |  | Female |  |
|  | Estimate* (95% CI) | P value | Estimate* (95% CI) | P value |
| Segmental airway wall thickness (mm) | 0.010 (0.004, 0.017) | 0.0008 | 0.010 (0.005, 0.014) | 0.0002 |
| Segmental airway lumen diameter (mm) | -0.052 (-0.111, 0.008) | 0.088 | -0.043 (-0.093, 0.007) | 0.091 |
| Subsegmental airway wall thickness (mm) | 0.009 (0.003, 0.015) | 0.0035 | 0.002 (-0.003, 0.007) | 0.42 |
| Subsegmental airway lumen diameter (mm) | -0.029 (-0.087, 0.029) | 0.33 | -0.005 (-0.055, 0.045) | 0.84 |
| FEV<sub>1</sub> percent predicted (%) | -2.80 (-3.99, -1.60) | <0.001 | -1.20 (-2.29, -0.12) | 0.030 |
| FEV<sub>1</sub>/FVC (%) | 0.10 (-0.42, 0.61) | 0.71 | 0.003 (-0.45, 0.45) | 0.99 |
| Wheezing | 1.17 (0.98, 1.41) | 0.079 | 1.15 (0.97, 1.37) | 0.10 |
| Cough | 0.96 (0.71, 1.29) | 0.77 | 1.04 (0.82, 1.32) | 0.73 |
| Dyspnea | 1.25 (0.97, 1.61) | 0.087 | 1.22 (1.02, 1.46) | 0.031 |

ITAT volume was standardized into sex-specific z-scores with a mean of 0 and a standard deviation of 1 within each sex.
Models included ITAT volume, waist circumference, age, current smoking status, smoking pack-years, height, and study cohort.
\* Linear regression coefficient ( $\beta$ ) for airway and spirometry outcomes, odds ratio for respiratory symptoms. Estimate was based on per 1-standard-deviation increase in ITAT volume or waist circumference.
FEV<sub>1</sub>, forced expiratory volume in 1 second; FVC, forced vital capacity; CI, confidence interval.

**Table S5.** Multivariable regression analyses of the associations between intrathoracic adipose tissue (ITAT) quintiles and airway-related outcomes.

| Outcome | Quintile | Effect measure | Estimate (95% CI) | P value |
| --- | --- | --- | --- | --- |
| Segmental wall thickness (mm) | Q1 | $\beta$ | Reference | |
| | Q2 | $\beta$ | 0.006 (-0.004, 0.015) | 0.24 |
| | Q3 | $\beta$ | 0.004 (-0.006, 0.014) | 0.42 |
| | Q4 | $\beta$ | 0.012 (0.002, 0.023) | 0.019 |
| | Q5 | $\beta$ | 0.026 (0.014, 0.038) | <0.0001 |
| Segmental lumen diameter (mm) | Q1 | $\beta$ | Reference | |
| | Q2 | $\beta$ | -0.075 (-0.166, 0.015) | 0.10 |
| | Q3 | $\beta$ | -0.161 (-0.257, -0.065) | 0.0010 |
| | Q4 | $\beta$ | -0.134 (-0.235, -0.033) | 0.0094 |
| | Q5 | $\beta$ | -0.099 (-0.215, 0.017) | 0.095 |
| Subsegmental wall thickness (mm) | Q1 | $\beta$ | Reference | |
| | Q2 | $\beta$ | 0.003 (-0.007, 0.012) | 0.58 |
| | Q3 | $\beta$ | 0.003 (-0.007, 0.012) | 0.60 |
| | Q4 | $\beta$ | 0.008 (-0.002, 0.018) | 0.13 |
| | Q5 | $\beta$ | 0.014 (0.002, 0.025) | 0.022 |
| Subsegmental lumen diameter (mm) | Q1 | $\beta$ | Reference | |
| | Q2 | $\beta$ | -0.057 (-0.145, 0.031) | 0.20 |
| | Q3 | $\beta$ | -0.045 (-0.139, 0.049) | 0.35 |
| | Q4 | $\beta$ | -0.016 (-0.115, 0.082) | 0.75 |
| | Q5 | $\beta$ | -0.016 (-0.130, 0.098) | 0.78 |
| FEV<sub>1</sub> percent predicted (%) | Q1 | $\beta$ | Reference | |
| | Q2 | $\beta$ | -0.77 (-2.60, 1.06) | 0.41 |
| | Q3 | $\beta$ | -1.34 (-3.31, 0.63) | 0.18 |
| | Q4 | $\beta$ | -2.18 (-4.28, -0.07) | 0.043 |
| | Q5 | $\beta$ | -4.80 (-7.21, -2.38) | 0.0001 |
| FEV<sub>1</sub>/FVC (%) | Q1 | $\beta$ | Reference | |
| | Q2 | $\beta$ | -1.01 (-1.78, -0.25) | 0.0097 |
| | Q3 | $\beta$ | -0.68 (-1.51, 0.14) | 0.11 |
| | Q4 | $\beta$ | -0.49 (-1.38, 0.39) | 0.27 |
| | Q5 | $\beta$ | -0.64 (-1.65, 0.38) | 0.22 |
| Wheezing | Q1 | OR | Reference |  |
|  | Q2 | OR | 1.34 (0.93, 1.94) | 0.12 |
|  | Q3 | OR | 1.71 (1.17, 2.48) | 0.0050 |
|  | Q4 | OR | 1.68 (1.14, 2.48) | 0.0091 |
|  | Q5 | OR | 2.06 (1.34, 3.15) | 0.0009 |
| Cough | Q1 | OR | Reference |  |
|  | Q2 | OR | 0.96 (0.57, 1.61) | 0.87 |
|  | Q3 | OR | 1.17 (0.69, 1.98) | 0.57 |
|  | Q4 | OR | 1.05 (0.61, 1.82) | 0.87 |
|  | Q5 | OR | 1.19 (0.65, 2.17) | 0.58 |
| Dyspnea | Q1 | OR | Reference |  |
|  | Q2 | OR | 1.08 (0.62, 1.88) | 0.80 |
|  | Q3 | OR | 1.14 (0.66, 1.98) | 0.63 |
|  | Q4 | OR | 1.36 (0.79, 2.34) | 0.26 |
|  | Q5 | OR | 1.83 (1.03, 3.24) | 0.039 |

Models included sex-specific ITAT quintiles, waist circumference, age, sex, current smoking status, smoking pack-years, height, and study cohort.
FEV<sub>1</sub>, forced expiratory volume in 1 second; FVC, forced vital capacity; CI, confidence interval; OR, odds ratio.

